# A systematic review of persistent symptoms and residual abnormal functioning following acute COVID-19: Ongoing symptomatic phase vs. post-COVID-19 syndrome

**DOI:** 10.1101/2021.06.25.21259372

**Authors:** Glenn Jennings, Ann Monaghan, Feng Xue, David Mockler, Román Romero-Ortuño

## Abstract

**Objective:** To compare the two phases of long COVID, namely ongoing symptomatic COVID-19 (OSC; signs and symptoms from 4 to 12 weeks from initial infection) and post-COVID-19 syndrome (PCS; signs and symptoms beyond 12 weeks) with respect to symptomatology, abnormal functioning, psychological burden, and quality of life.

**Design:** Systematic review.

**Data Sources:** Electronic search of EMBASE, MEDLINE, ProQuest Coronavirus Research Database, LitCOVID, and Google Scholar between January and April 2021, and manual search for relevant citations from review articles.

**Eligibility Criteria:** Cross-sectional studies, cohort studies, randomised control trials, and case-control studies with participant data concerning long COVID symptomatology or abnormal functioning.

**Data Extraction:** Studies were screened and assessed for risk of bias by two independent reviewers, with conflicts resolved with a third reviewer. The AXIS tool was utilised to appraise the quality of the evidence. Data were extracted and collated using a data extraction tool in Microsoft Excel.

**Results:** Of the 1,145 studies screened, 39 were included, all describing adult cohorts with long COVID and sample sizes ranging from 32 to 1,733. Studies included data pertaining to symptomatology, pulmonary functioning, chest imaging, cognitive functioning, psychological disorder, and/or quality of life. Fatigue presented as the most prevalent symptom during both OSC and PCS at 43% and 44%, respectively. Sleep disorder (36%; 33%), dyspnoea (31%; 40%), and cough (26%; 22%) followed in prevalence. Abnormal spirometry (FEV_1_ <80% predicted) was observed in 15% and 11%, and abnormal chest imaging observed in 34% and 28%, respectively. Cognitive impairments were also evident (20%; 15%), as well as anxiety (28%; 34%) and depression (25%; 32%). Decreased quality of life was reported by 40% of patients with OSC and 57% by those with PCS.

**Conclusions:** The prevalences of OSC and PCS were highly variable. Reported symptoms covered a wide range of body systems, with general overlap in frequencies between the two phases. However, abnormalities in lung function and imaging seemed to be more common in OSC, whilst anxiety, depression, and poor quality of life seemed more frequent in PCS. In general, the quality of the evidence was moderate and further research is needed to better understand the complex interplay of somatic versus psychosocial drivers in long COVID.

**Systematic Review Registration:** Registered with PROSPERO with ID #CRD42021247846.

## 1 INTRODUCTION

On 11^th^ March 2020, the World Health Organisation (WHO) Director-General declared the COVID-19 outbreak a global pandemic (1) and, at the time of writing, over 175 million positive cases and almost 4 million deaths have been confirmed worldwide (2). Caused by the novel severe acute respiratory syndrome coronavirus 2 (SARS-CoV-2), COVID-19 represents a highly heterogenous disease affecting the respiratory tract and multiple other organ systems, with fever, fatigue, and cough presenting as the most prevalent symptoms (3). Less commonly reported symptoms include hyposmia, dyspnoea, headache, sore throat, and dizziness. Severity of COVID-19 manifestations ranges from asymptomatic to severe, with acute presentations often requiring invasive ventilation or extended stays in intensive care for patients (4). Overall, the acute COVID-19 phase typically endures for a period of up to 4 weeks from the onset of initial infection (5). In a subset of patients, symptoms can persist beyond the 4-week acute COVID-19 period into a post-acute phase that has been coined ‘long COVID’ (5). Long COVID can be further distinguished as ‘ongoing symptomatic COVID-19’ (OSC) and ‘post-COVID-19 syndrome’ (PCS), terms that describe persistent signs and/or symptoms in the periods from 4 to 12 weeks and over 12 weeks post-infection onset, respectively (5).

Due to the recentness of the COVID-19 pandemic, and the initial focus of research being on the acute phase symptomatology and treatment, an accurate characterisation of long COVID symptomatology in its distinct phases has remained elusive (6). Addressing the gap could help the understanding of long COVID predictors and help clinicians enhance symptom management and treatment. Thus, in this systematic review, we aimed to characterise and compare the OSC and OCS phases of long COVID, with an emphasis on prevalence, symptomatology, pulmonary and cognitive functioning, mental health aspects, and quality of life.

## 2 METHODS AND MATERIALS

### 2.1 Protocol registration

The review protocol was registered with PROSPERO, the international prospective register of systematic reviews by the National Institute of Health Research (ID: CRD42021247846). The protocol can be accessed on the PROSPERO register (7).

### 2.2 Search strategy

A search strategy was created by a medical librarian that included MeSH terminology related to “post-acute COVID-19”, “long COVID”, “prevalence”, “symptomatology”, “spirometry”, “imaging”, “cognitive”, “psychological burden”, and “quality of life”. The full search strategy is shown in Appendix A. EMBASE, MEDLINE, ProQuest Coronavirus Research Database, LitCOVID, and Google Scholar were searched between January and April 2021, with the search was limited to articles published between March 2020 and April 2021. A manual search of review articles’ reference lists was also conducted to identify relevant citations.

### 2.3 Eligibility criteria and study selection

Studies with samples sizes of 30 or more participants aged at least 18 years old reporting data on long COVID symptomatology and/or general post-acute COVID-19 functioning were included in the review. In terms of study designs, cross-sectional studies, cohort studies, randomised control trials, and case-control studies were included, while meta-analyses, systematic reviews, narrative reviews, clinical trials, case studies and series, opinion pieces, and non-peer reviewed publications were excluded. Studies with a gender imbalance greater than 80:20% in either direction were also excluded, as well as those reporting on specific cohorts (e.g. only patients with anosmia). Table 1 summarises the full eligibility criteria.

**Table 1.** Eligibility Criteria for Studies and Participants.

|  | Inclusion Criteria | Exclusion Criteria |
| --- | --- | --- |
| **Study Topic** | Studies with participant data concerning long COVID symptomatology and/or general post-acute COVID-19 functioning. | N/A |
| **Study Design** | Cross-sectional studies, cohort studies, randomised control trials, and case-control studies. | Meta-analyses, systematic reviews, narrative reviews, clinical trials, case studies and series, opinion pieces, and non-peer reviewed publications. |
| **Condition of Participants** | Participants who tested positive for SARS-CoV-2 infection or were suspected of SARS-CoV-2 infection. | Participants recovered from acute COVID-19 (denoted as $\geq 4$ weeks following symptom onset or hospital admission; immediately following discharge from hospital; or indicated as "recovered" by the respective researchers). |
| **Sample Size** | N/A | Studies with less than 30 participants. |
| **Participant Age** | N/A | Participants younger than 18 years of age |
| **Participant Gender** | N/A | Studies with a gender imbalance greater than 80:20%. |
| **Other** | N/A | Entire participant cohorts with a specific characteristic (e.g. only patients with anosmia). |

Citations generated from the search strategy were imported into a systematic review management tool, Covidence [covidence.org]. All duplicate imports were removed and an initial screening was conducted by two independent reviewers, with conflicts resolved with a third reviewer. All texts were then further screened by a single reviewer and studies adhering to the inclusion criteria were included in the data extraction stage. Studies were selected in accordance with the PICOS framework (Participants, Interventions, Comparisons, Outcomes, and Study Design) based on the Preferred Reporting Items for Systematic Reviews and Meta-Analyses (PRISMA) guidelines (8).

### 2.4 Data extraction

The data from the included studies were extracted by a single reviewer using Microsoft Excel (Appendix B). Data included were as follows: *i*) study details (i.e. first author, date of publication, country of authorship, topic of the study, and study design); *ii*) population details (i.e. sample size, mean/median age, gender proportion, eligibility criteria, acute COVID-19 hospitalisation status, and time post-COVID-19 onset); *iii*) prevalence data of residual symptoms; and *iv*) prevalence data of abnormal cognitive, pulmonary, and chest imaging findings, and poor mental health and quality of life data. Missing data were requested from the respective corresponding authors, if necessary.

The timepoints of assessment were adjusted for uniformity, with ‘time’ relating to the number of weeks following initial onset of acute COVID-19. For studies that reported time following acute phase recovery or hospital discharge, a 4-week acute phase period was inserted in accordance with NICE guidelines (5). The clinical data were then recorded as individual prevalences at single timepoints, with several prevalence points collected in longitudinal studies. Prevalences within 4-12 weeks and after 12 weeks were collated to produce a mean (+range) prevalence per symptom in the OSC and PCS phases, respectively. An overarching long COVID prevalence incorporating all the data per symptom was also calculated. Prevalence data were only recorded for either OSC and PCS in symptoms or abnormal traits identified at three or more distinct assessment timepoints. The entire data synthesis strategy was completed via Microsoft Excel.

### 2.5 Quality appraisal and risk of bias

The AXIS Critical Appraisal Tool (9) was applied to each included study by two independent reviewers. For each study, a score out of 20 was generated and any disparities were resolved with a third reviewer.

## 3 RESULTS

### 3.1 Description of included studies

A total of 1,445 studies were retrieved from the online databases, with a further 37 identified through references of review articles. After 292 duplicates were removed, an initial screening of the remaining 1,190 studies was conducted. 179 studies were included for further screening which produced a final list of 39 studies for data extraction (10-48). A PRISMA flow diagram outlining the screening process is provided in Figure 1.

**Figure 1.**
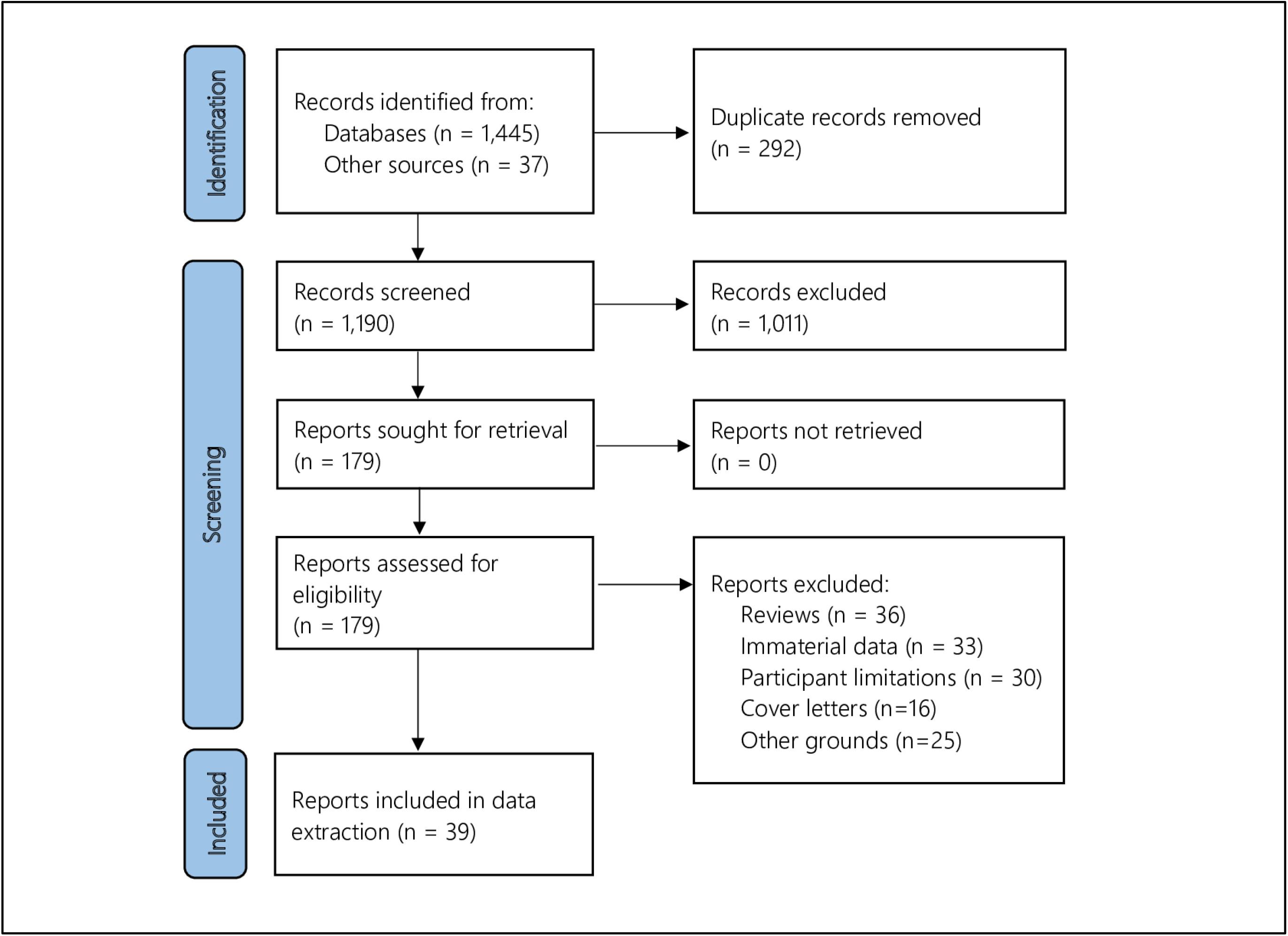
PRISMA flow diagram.

The main characteristics of the 39 included studies are presented in Table 2. Studies were conducted in 17 different countries. The sample sizes ranged from 32 to 1,733, whilst participants’ ages ranged from 32 to 74 years and proportions of female participants between 31% and 72%. Participants’ hospitalisation status varied between the studies, with 69% (n = 27), 3% (n = 1), and 28% (n = 11) addressing inpatient, non-hospitalised, and mixed cohorts, respectively. Assessment time post-COVID-19 onset was between 4 and 31 weeks, with data available at 51 timepoints: 29 during OSC and 22 during PCS.

**Table 2.**
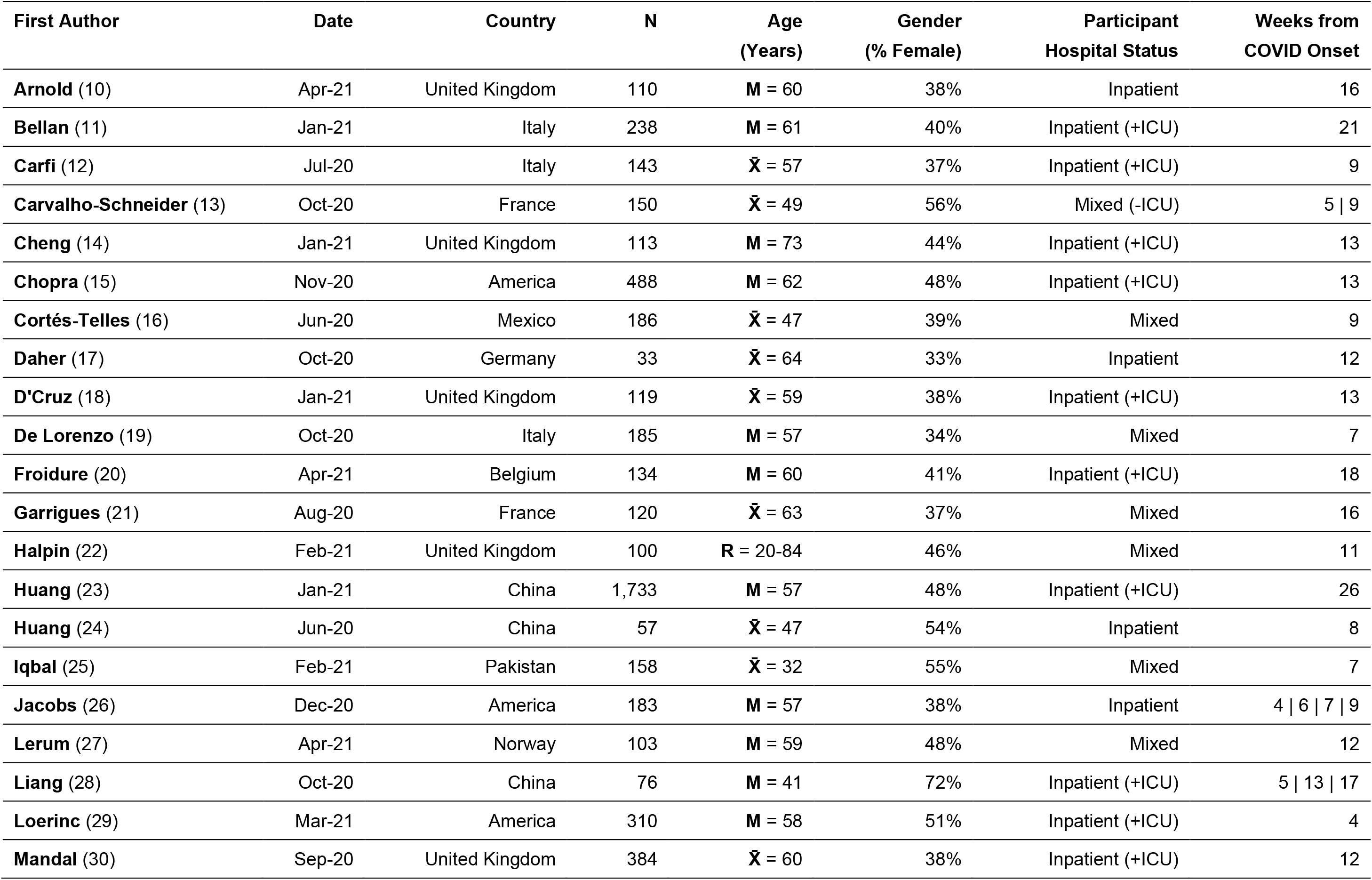
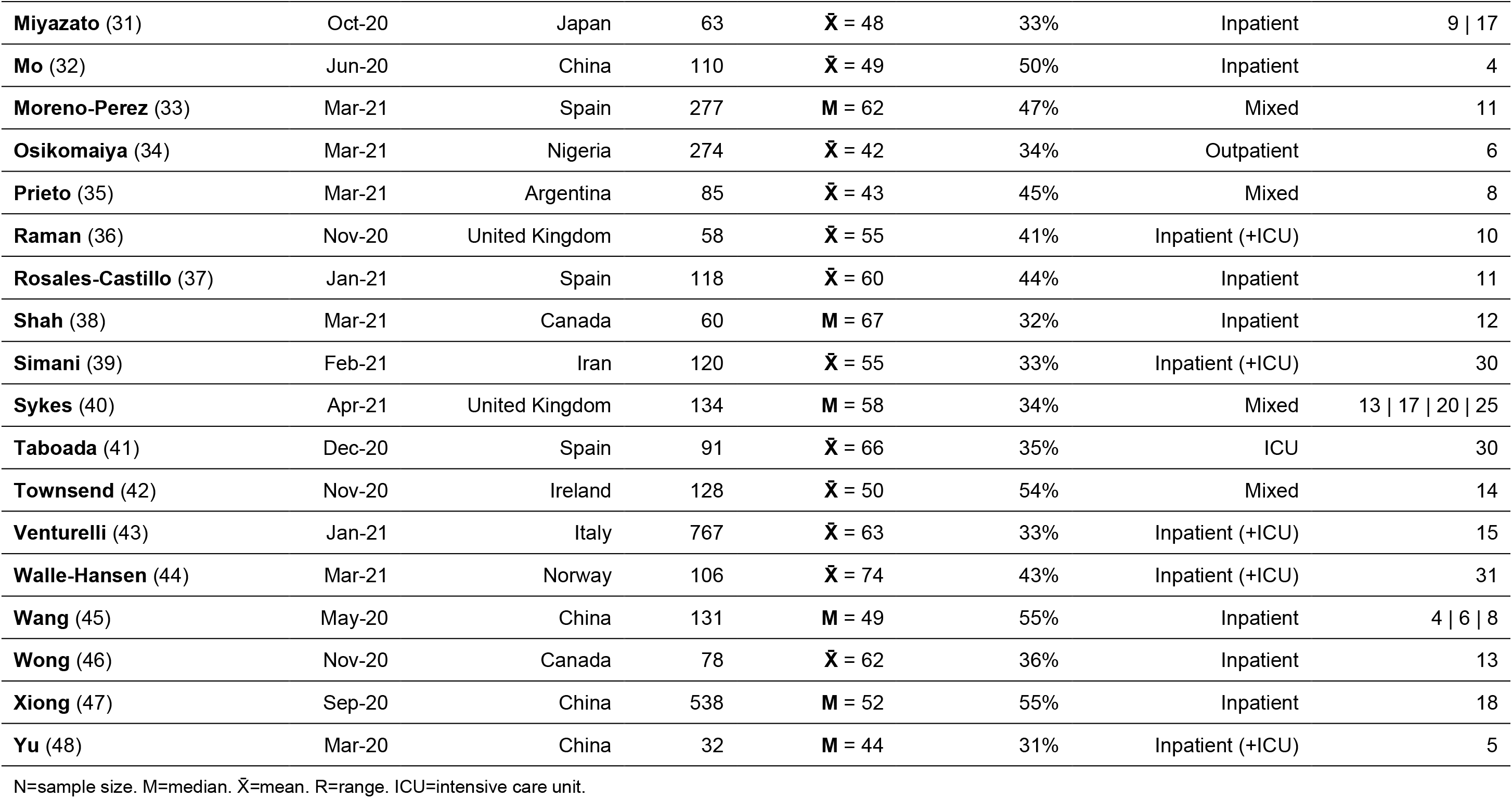
Demographics of Included Studies.

### 3.2 Quality appraisal and risk of bias

The average AXIS score for all included studies was 16.9 (±2.0) out of a possible 20, which may indicate a moderate risk of bias. The major sources of bias were the use of the convenience sampling methods, which was utilised by 38 of the 39 studies (10-24, 26-49), and possible non-response bias in 12 studies (16, 17, 25-27, 32, 34-39, 48). The results of the AXIS critical appraisal for each included study are displayed in Table 3.

**Table 3.**
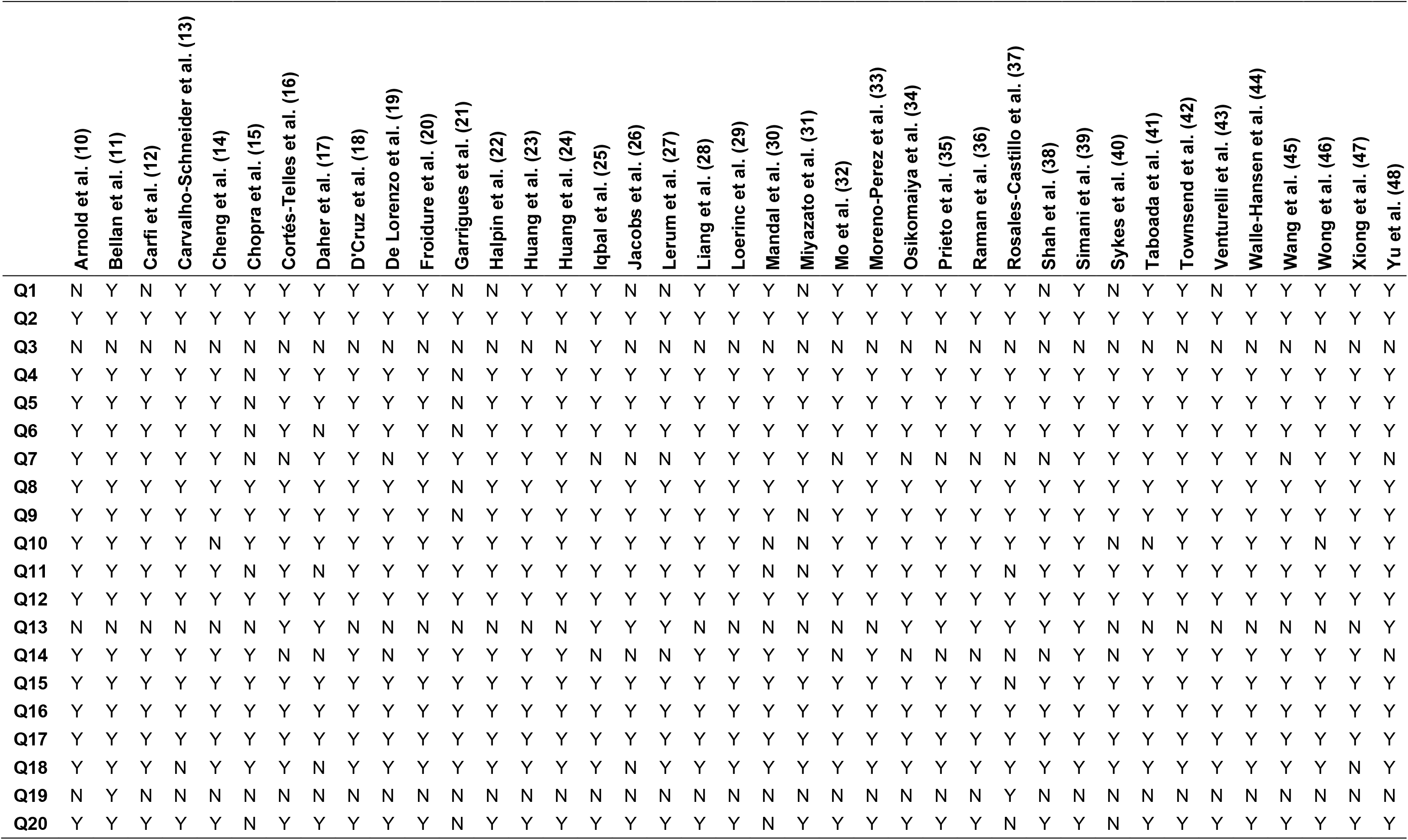
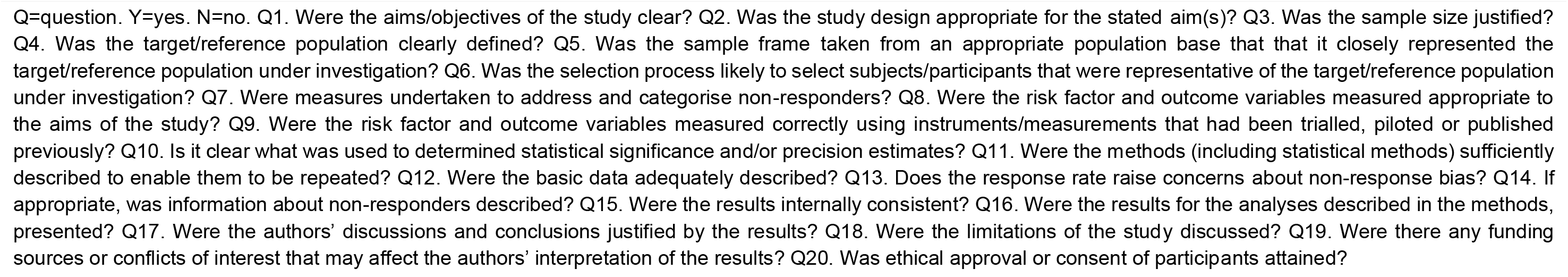
AXIS Critical Appraisal.

### 3.3 Ongoing symptomatic COVID-19 and post-COVID-19 syndrome

Based on NICE criteria (5), the diagnoses of OSC or PCS were denoted by prevalence of at least one persistent symptom or sign. Overall, the presence of one or more symptoms in patients was recorded from 20 studies during long COVID (10, 12-15, 18, 23, 26, 30, 33-35, 37, 39, 41-43, 45-47), with two studies presenting longitudinal data (13, 45). OSC was recorded in 9 distinct studies, with a mean prevalence of 59% and a range from 14% to 87%. As for PCS, a prevalence of 62% for at least one symptom was identified from a total of 11 studies, with a range between 18% and 89%. Figure 2 depicts the reported prevalences of these two long COVID phases.

**Figure 2.**
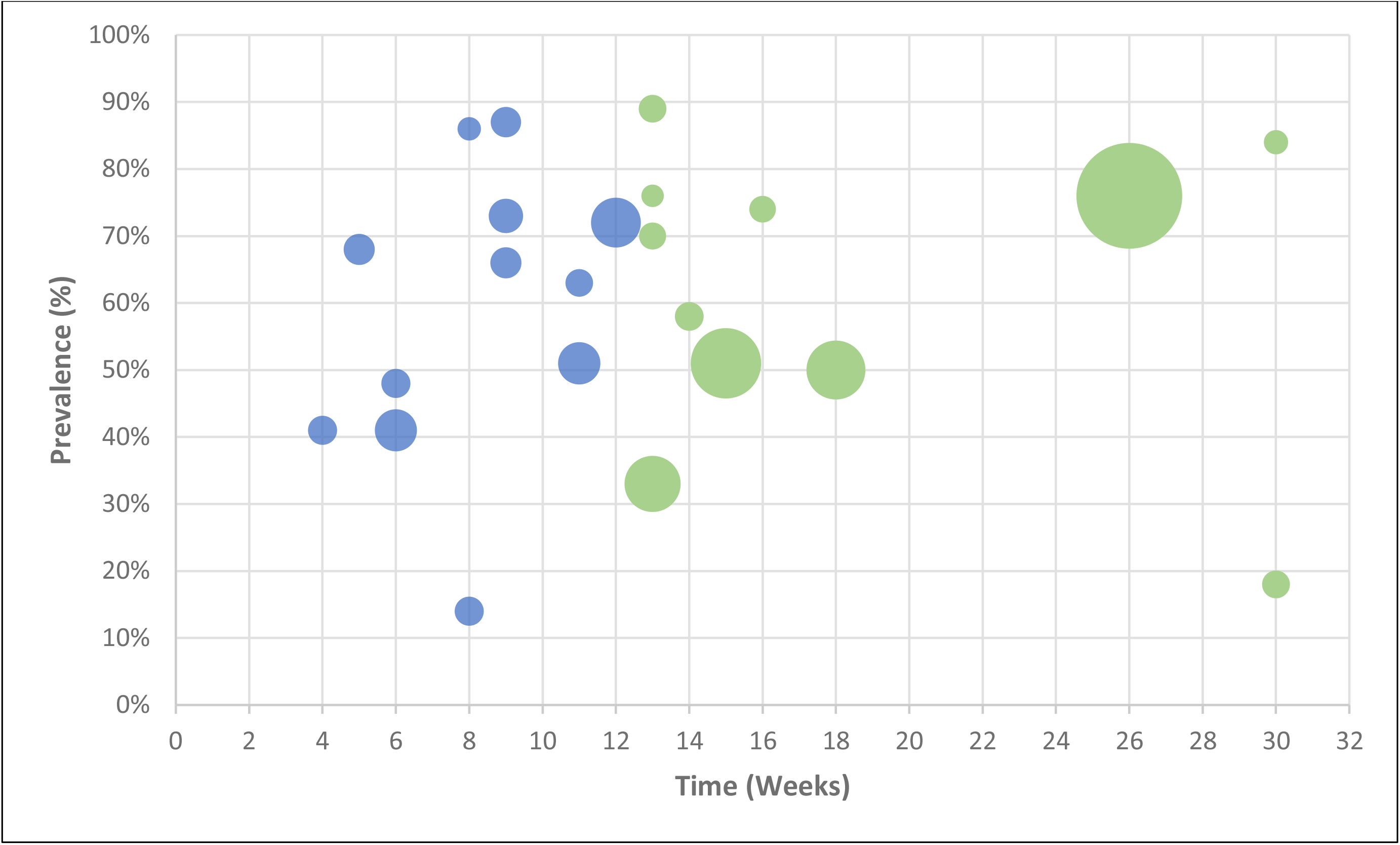
Bubble chart of the reported prevalences of the two long COVID phases (ongoing symptomatic COVID-19 in blue; post-COVID-19 syndrome in green), where the size of each bubble is proportional to the study sample size.

### 3.4 Symptomatology

Figure 3 provides an overview of the mean prevalence proportions of OCS and PSC symptoms across body symptoms, and Table 4 details the prevalence ranges and number of assessment timepoints involved.

**Figure 3.**
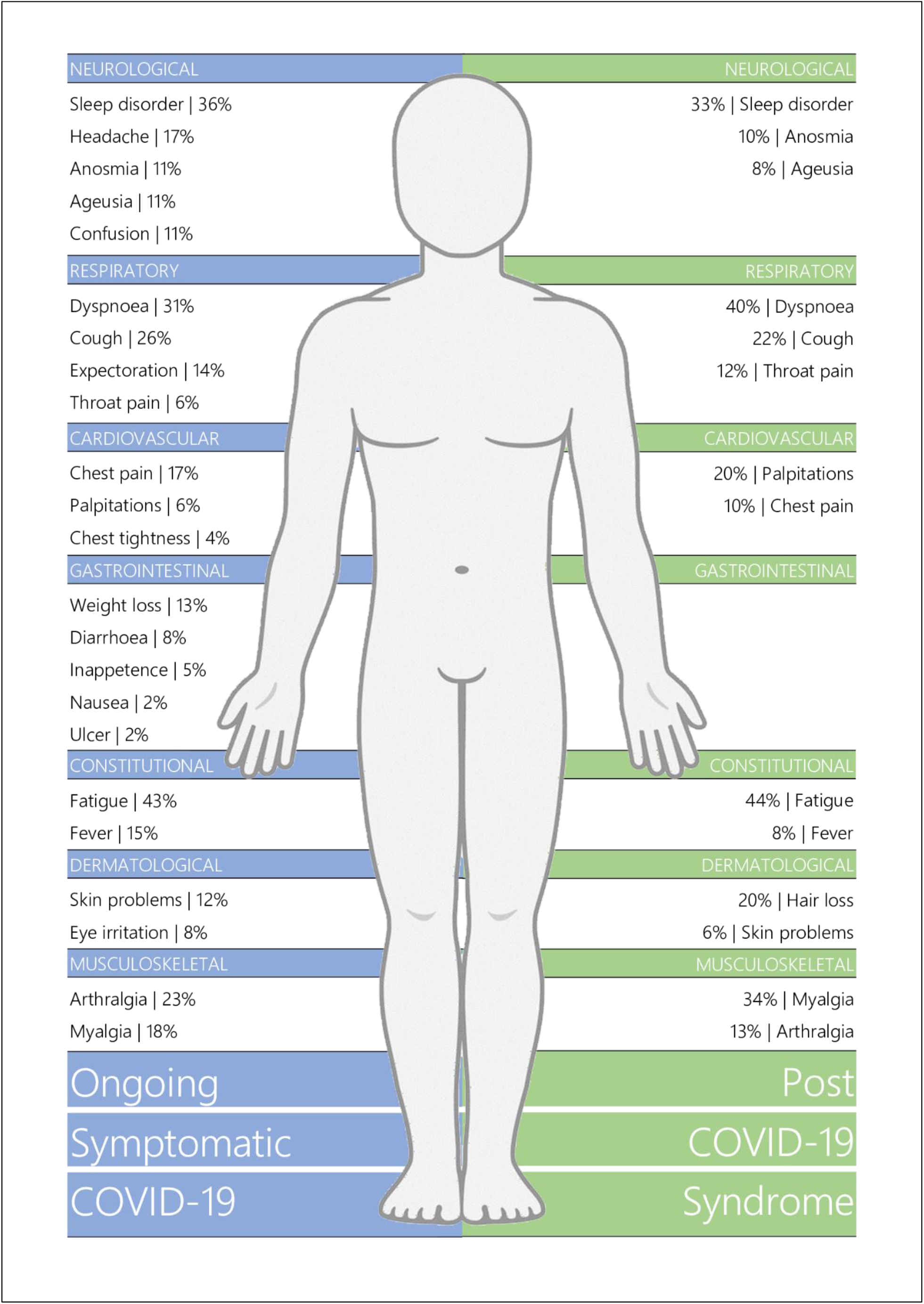
Body Chart of Long COVID Symptomatology.

**Table 4.** Symptom Prevalence of Long COVID Patients.

|  | Ongoing Symptomatic COVID-19 |  |  |  |  | Post-COVID-19 Syndrome |  |  |  |  |
| --- | --- | --- | --- | --- | --- | --- | --- | --- | --- | --- |
| | $\bar{x}$ | SD | N | Min. | Max. | $\bar{x}$ | SD | N | Min. | Max. |
| **Constitutional** |  |  |  |  |  |  |  |  |  |  |
| Fatigue | 43% | 24 | 19 | 5% | 83% | 44% | 19 | 16 | 10% | 71% |
| Fever | 14% | 18 | 8 | 1% | 51% | 8% | 8 | 7 | 1% | 20% |
| **Respiratory** |  |  |  |  |  |  |  |  |  |  |
| Dyspnoea | 31% | 19 | 25 | 2% | 64% | 40% | 21 | 15 | 6% | 73% |
| Cough | 26% | 13 | 19 | 5% | 45% | 22% | 16 | 16 | 3% | 59% |
| Expectoration | 13% | 8 | 7 | 1% | 25% | - | - | - | - | - |
| Throat pain | 6% | 6 | 7 | 1% | 17% | 12% | 9 | 6 | 3% | 29% |
| **Neurological** |  |  |  |  |  |  |  |  |  |  |
| Sleep disorder | 36% | 25 | 5 | 10% | 69% | 33% | 13 | 11 | 18% | 57% |
| Headache | 17% | 8 | 10 | 4% | 36% | - | - | - | - | - |
| Anosmia | 11% | 7 | 9 | 2% | 21% | 10% | 3 | 8 | 5% | 13% |
| Ageusia | 11% | 9 | 8 | 1% | 25% | 8% | 4 | 7 | 2% | 15% |
| Confusion | 11% | 3 | 3 | 9% | 14% | - | - | - | - | - |
| **Cardiovascular** |  |  |  |  |  |  |  |  |  |  |
| Chest pain | 17% | 11 | 9 | 3% | 35% | 10% | 6 | 11 | 1% | 22% |
| Palpitations | 6% | 4 | 5 | 2% | 11% | 20% | 28 | 4 | 4% | 62% |
| Chest tightness | 4% | 3 | 3 | 1% | 6% | - | - | - | - | - |
| **Gastrointestinal** |  |  |  |  |  |  |  |  |  |  |
| Weight loss | 13% | 6 | 3 | 6% | 17% | - | - | - | - | - |
| Diarrhoea | 8% | 5 | 10 | 1% | 18% | - | - | - | - | - |
| Inappetence | 5% | 4 | 4 | 1% | 9% | - | - | - | - | - |
| Nausea | 2% | 2 | 5 | 1% | 6% | - | - | - | - | - |
| Ulcer | 2% | 1 | 3 | 1% | 3% | - | - | - | - | - |
| **Musculoskeletal** |  |  |  |  |  |  |  |  |  |  |
| Arthralgia | 23% | 13 | 7 | 10% | 48% | 13% | 11 | 4 | 6% | 29% |
| Myalgia | 18% | 10 | 9 | 1% | 32% | 34% | 31 | 9 | 2% | 86% |
| **Dermatological** |  |  |  |  |  |  |  |  |  |  |
| Skin problems | 12% | 4 | 3 | 8% | 15% | 6% | 4 | 4 | 3% | 12% |
| Eye irritation | 8% | 3 | 4 | 4% | 11% | - | - | - | - | - |
| Hair loss | - | - | - | - | - | 20% | 9 | 5 | 6% | 29% |

$\bar{x}$ =mean. SD=standard deviation. N=number of assessment timepoints. Min.=minimum. Max.=maximum.

#### 3.4.1 Ongoing symptomatic COVID-19

The most prevalent symptom in patients with OSC was fatigue with a mean prevalence of 43% (range: 5-83%). Sleep disorders were also highly prevalent at 36% (10-69%), with dyspnoea (31%; 2-64%) and cough (26%; 5-45%) reported as the most common respiratory symptoms. Other symptoms identified in patients between 4-12 weeks included arthralgia (23%; 10-48%), myalgia (18%; 1-32%), chest pain (17%; 3-35%), headache (17%; 4-36%), fever (15%; 1-51%), expectoration (14%; 1-25%), weight loss (13%; 6-17%), skin problems (12%; 8-15%), anosmia (11%; 2-21%), ageusia (11%; 1-25%), and confusion (11%; 9-14%). Less common manifestations were eye irritation (8%; 4-11%), diarrhoea (8%; 1-18%), throat pain (6%; 1-17%), palpitations (6%; 2-11%), inappetence (5%; 1-9%), chest tightness (4%; 1-6%), nausea (2%; 1-6%), and peptic ulcer (2%; 1-3%).

#### 3.4.2 Post-COVID-19 syndrome

Fatigue also presented as the most common symptom in PCS patients at 44% (10-71%), with dyspnoea, myalgia, and sleep disorder also highly prevalent at a mean of 40% (6-73%), 34% (2-86%), and 33% (18-57%), respectively. Other symptoms reported in patients over 12 weeks post-disease onset included cough (22%; 3-59%), hair loss (20%; 6-29%), palpitations (20%; 4-62%), arthralgia (13%; 6-29%), throat pain (12%; 3-29%), anosmia (10%; 5-13%), and chest pain (10%; 1-22%). Fever (8%; 1-20%), ageusia (8%; 2-15%), and skin problems (6%; 3-12%) were less commonly reported.

### 3.5 Respiratory functioning

#### 3.5.1 Pulmonary functioning

Table 5 summarises the prevalence of abnormal pulmonary function parameters across included studies, which include forced expiratory volume in the first second (FEV_1_), forced vital capacity (FVC), the FEV_1_/FVC ratio, and diffusion capacity for carbon monoxide (DL_CO_). During the OSC phase, FEV_1_ values below of the predicted normal were identified in a mean of 15% (9-21%) of patients. Abnormal FVC scores averaged a prevalence of 12% (7-21%), and FEV_1_/FVC and DL_CO_ impairments were identified in 6% (1-11%) and 44% (24-53%) of patients, respectively. During the PSC phase, the mean prevalence of abnormal FEV_1_ was 11% (5-17%), and those of FVC, FE1/FVC ratio, and DLCO were 11% (1-19%), 7% (6-8%), and 32% (20-46%), respectively.

**Table 5.** Prevalence of Pulmonary and Cognitive Functioning, Psychological Burden, and Quality of Life.

|  | Ongoing Symptomatic COVID-19 |  |  |  |  | Post-COVID-19 Syndrome |  |  |  |  |
| --- | --- | --- | --- | --- | --- | --- | --- | --- | --- | --- |
| | $\bar{x}$ | SD | N | Min. | Max. | $\bar{x}$ | SD | N | Min. | Max. |
| **Pulmonary Functioning** |  |  |  |  |  |  |  |  |  |  |
| FEV<sub>1</sub> < 80% predicted | 15% | 5 | 5 | 9% | 21% | 11% | 6 | 4 | 5% | 17% |
| FVC < 80% predicted | 12% | 5 | 5 | 7% | 21% | 11% | 9 | 4 | 1% | 19% |
| FEV<sub>1</sub>/FVC < 0.7 | 6% | 4 | 4 | 1% | 11% | 7% | 1 | 3 | 6% | 8% |
| DL<sub>CO</sub> < 80% predicted | 44% | 14 | 4 | 24% | 53% | 32% | 11 | 4 | 20% | 46% |
| **Chest Imaging** |  |  |  |  |  |  |  |  |  |  |
| Abnormal pattern(s) | 34% | 25 | 5 | 2% | 60% | 28% | 17 | 5 | 13% | 53% |
| Ground-glass opacity | 28% | 29 | 3 | 1% | 59% | 24% | 26 | 6 | 2% | 67% |
| Fibrosis | 19% | 22 | 3 | 5% | 44% | 7% | 9 | 4 | 2% | 20% |
| Reticulation | - | - | - | - | - | 11% | 12 | 3 | 1% | 24% |
| Consolidation | - | - | - | - | - | 3% | 3 | 3 | 1% | 7% |
| **Cognitive Impairments** |  |  |  |  |  |  |  |  |  |  |
| Cognitive impairment | 20% | 11 | 5 | 2% | 28% | 15% | 6 | 5 | 5% | 22% |
| Concentration issues / Attention issues | - | - | - | - | - | 30% | 9 | 5 | 21% | 43% |
| Memory impairment | - | - | - | - | - | 35% | 16 | 6 | 6% | 48% |
| **Psychological Disorder** |  |  |  |  |  |  |  |  |  |  |
| Anxiety | 28% | 18 | 4 | 14% | 53% | 34% | 21 | 8 | 6% | 62% |
| Depression | 25% | 15 | 3 | 15% | 42% | 32% | 24 | 9 | 4% | 76% |
| Post-traumatic stress | - | - | - | - | - | 18% | 12 | 3 | 6% | 31% |
| **Quality of Life** |  |  |  |  |  |  |  |  |  |  |
| Decreased quality of life | 40% | 15 | 3 | 23% | 53% | 57% | 9 | 3 | 51% | 67% |
| Decrease in usual activities | - | - | - | - | - | 23% | 17 | 4 | 2% | 37% |
| Mobility issues | 51% | 15 | 3 | 37% | 67% | 32% | 25 | 3 | 7% | 56% |
| Pain or discomfort | - | - | - | - | - | 36% | 11 | 3 | 27% | 48% |
| Depression / Anxiety | - | - | - | - | - | 27% | 14 | 4 | 14% | 46% |
| Issues with self-care | - | - | - | - | - | 10% | 7 | 4 | 1% | 17% |

$\bar{x}$ =mean. SD=standard deviation. N= number of assessment timepoints. Min.=minimum. Max.=maximum. FEV<sub>1</sub>=forced expiratory volume in one second. FVC=forced vital capacity. DL<sub>CO</sub>=lung diffusion capacity for carbon monoxide.

#### 3.5.2 Lung imaging

Lung imaging was performed at 15 assessment points using computed tomography (CT; n=6), high-resolution CT (HRCT; n=6), chest radiography (CXR; n=5), and/or magnetic resonance imaging (MRI; n=1). Overall, abnormal imaging patterns were observed in 34% (2-60%) of patients with OSC, with specific abnormalities including ground-glass opacity (28%; 1-59%) and fibrosis (19%; 5-44%) (Table 5). During the PCS phase, a prevalence of 28% (13-53%) was identified for abnormal patterns; ground-glass opacity was the most prevalent abnormality at 24% (2-67%), with reticulation (11%; 1-24%), fibrosis (7%; 2-20%), and consolidation (3%; 1-7%) also recorded in a subset of patients (Table 5).

### 3.6 Cognitive functioning

Data on cognitive impairments were available at both phases of long COVID from a total of 10 distinct timepoints (17-19, 22, 33, 36, 40). Data regarding specific modalities of cognition such as memory, concentration, and attention were available for PCS studies only (14, 21, 22, 33, 40) and are presented in Table 5. A mean proportion of 20% (2-28%) of patients were reported to have cognitive impairment during the OSC phase, and 15% (5-22%) during PCS. Both concentration or attention issues and memory deficits were prevalent at 30% (21-43%) and 35% (6-48%), respectively, in patients with PCS.

### 3.7 Mental health & quality of life

During the OSC phase, anxiety and depression were reported in a mean of 28% (14-53%) and 25% (15-42%), respectively (Table 5). 40% (23-53%) of patients also expressed a decreased quality of life. The EQ-5D-5L was utilised to assess quality of life data, with this measure incorporating sub-scales to explore five dimensions of quality of life (50). Mobility issues were reported in a mean of 51% (37-67%) of patients who completed the EQ-5D-5L assessment during OSC, with insufficient data available for the remaining dimensions.

The PCS phase seemed to have higher mean prevalences of anxiety (34%; 6-62%) and depression (32%; 4-76%), whilst post-traumatic stress was also prevalent in 18% (6-31%) of patients. A decreased quality of life was recorded in 57% (51-67%) of the samples, with the EQ-5D-5L sub-scales identifying the following prevalence proportions: pain or discomfort (36%; 27-48%), mobility issues (32%; 7-56%), depression or anxiety (27%; 14-46%), a decrease in usual activities (23%; 2-37%), and issues with self-care (10%; 1-17%) (Table 5).

## 4 DISCUSSION

### 4.1 Statement of principal findings

The aim of this systematic review was to compare the two phases of long COVID, namely OSC (signs and symptoms from 4 to 12 weeks since initial infection) and PCS (signs and symptoms beyond 12 weeks), with respect to symptomatology, abnormal cognitive and respiratory functioning, psychological burden, and quality of life. Overall, findings indicate that the prevalence proportions of OSC and PCS were highly variable across studies, reflecting the non-probabilistic sampling of included studies and differences in hospitalisation status. Reported symptoms covered a wide range of body systems, with general overlap in frequency ranges between the two long COVID phases. Fatigue and sleep disorders seemed to have comparably high prevalences. Symptoms such as arthralgia, fever, and chest pain appeared to decrease with time, whilst myalgia, palpitations, and dyspnoea seemed to increase during the PCS phase. Data on expectoration, chest tightness, headache, confusion, gastrointestinal issues, and eye irritation was only available for the OSC phase (13, 16, 17, 22, 25, 26, 28, 33, 34, 45), whereas hair loss was only reported in patients with PCS (14, 21, 23, 31, 47). In terms of cognitive impairment, there seemed to be a slight mean prevalence reduction in the PCS phase, with specific data on concentration, attention, and memory being unavailable for the initial long COVID phase. Even though they also had overlapping frequencies, abnormalities in lung function and imaging seemed to have higher frequencies in OSC, whilst anxiety, depression, and poor quality of life seemed more frequent in PCS. Post-traumatic stress was only mentioned in PCS studies (11, 39, 43).

Overall, findings would suggest that OSC and PCS are a disease continuum with marked clinical overlap as opposed to discrete, easily distinguishable phases. However, results suggest the possibility that OSC may have a higher burden of somatic disease, while PCS may be characterised by a relatively higher psychosocial burden. However, in general, the quality of the evidence was moderate, and many symptoms were only reported in a subset of patients. Therefore, further research is needed to better understand the complex interplay between somatic and psychosocial drivers in long COVID.

### 4.2 Strengths and weaknesses of the study

A strength of the study is the novel approach to the characterisation of long COVID by considering the OSC and PCS phases, which NICE separated as potentially distinct entities (5) but had not yet been systematically characterised. Another robust aspect of this review is the collation of a total of 39 studies conducted in 17 different countries, which captures the global nature of the COVID-19 pandemic.

However, the major limitation of the study resides in the lack of inter-study consistency regarding assessment methods for symptomatology and functional impairments. Many of the studies denoted symptom presence or absence using self-report tools, which are affected by self-report biases (51). Standardised scales were also utilised, however there was no consistency in the selected scales with fatigue alone quantified by 5 distinct objective scales: the Chalder Fatigue Scale (42), the Fatigue Severity Scale (36), the PROMIS (26), and SF-36 (10) scales, and a previously validated unnamed scale (39). This poor inter-study consistency may compromise the validity of the findings, with scales potentially being more or less sensitive or even assessing distinct sub-domains of a symptom. Abnormal patterns in chest imaging were also highly heterogenous through the mixed use of chest x-ray, regular CT, high-resolution CT, and magnetic resonance imaging. Due to the limited data available, differences in assessment tools were not addressed in the eligibility screening phase of the review. Overall, the lack of inter-study consistency in methodology may explain the large ranges observed in the data. The moderate quality of the data acquired from the included studies must also be acknowledged in relation to the wide-ranging prevalence findings. An average AXIS score of 16.9 (±2.0) for the studies suggests that the results should be interpreted with caution (9).

### 4.3 Strengths and weaknesses in relation to other studies

Although the number of reviews attempting to characterise long COVID is exponentially increasing (49, 52-56), many of those published present a narrative, rather than systematic, discussion of the findings. In addition to adding value by characterising long COVID separately by OSC and PCS phases, our study offers a structured systematic overview of the long-term effects of COVID-19.

Another point of note regarding the present review is the inclusion of multisystem-related symptoms and impairments. While, previous reviews have focused solely on neurological or respiratory functioning (57-60), our review provides a more comprehensive and collective characterisation of long COVID and further evidences its heterogenous nature. We acknowledge, however, that our review is not fully comprehensive. For example, Nalbandian et al. (52) narratively described haematologic, renal, and endocrine post-acute COVID-19 complications, and these body systems were not incorporated into the present review’s literature search. Another potential limitation of the current review was the fact that patient hospitalisation status or acute phase history were not taken into account when characterising the signs and symptoms of OSC and PCS. While primary data for these characteristics were indeed presented by several studies (21, 27, 40), there were insufficient data available to provide a comprehensive distinction of patients’ outcomes with respect to them.

### 4.4 Meaning of the study

This systematic review provides clinicians, other healthcare professionals, and policymakers with a comprehensive, yet concise, characterisation of the two phases of long COVID, namely OSC and PCS. Overall, the findings provide a systematic description of the typical clinical profile of long COVID patients and could enhance the understanding of the condition, thereby potentially resulting in better treatment options and management of symptoms, and implementation of policies that allow long COVID patients to receive the best possible care. The suggested higher relative importance of psychosocial drivers in the PCS phase may inform more holistic assessment and treatment strategies, including psychological and psychosocial supports. Additionally, the frequent presence of psychological distress may be linked to several reported symptoms, with a range of psychological disorders often associated with hair loss (61), sleep disorders (62), gastrointestinal issues (63), pain (64), and cardiovascular symptoms (64). Establishing potential associations will further enhance patient care by enabling to cluster signs and symptoms, and characterise several ‘subtypes’ of long COVID.

### 4.5 Unanswered questions and future research

The suggested finding of this review is that the longer long COVID persists, the more important psychological and psychosocial aspects could be. With this, further research is needed to better establish the relative contributions of somatic versus psychosocial drivers in long COVID over time. Should research prove psychosocial drivers more important, this might help avoid inappropriate medicalisation of this still poorly understood condition and divert support resources where they are most needed (e.g. social security, psychological support). In addition, further characterisation is needed regarding the potential impact of acute phase presentation, hospitalisation status, age, sex, education, socioeconomic status, occupation, and baseline physical and psychological/psychiatric comorbidities on the risk of developing long COVID. In addition, there is scope for future studies linking long COVID clinical profiles to respective physiological and immunological profiles, to see whether or not they align in the pathophysiology of long COVID.

## Data Availability

The authors confirm that the data supporting the findings of this study are available within the article and its supplementary materials.

