## Appendix A for "A systematic review of persistent symptoms and residual abnormal functioning following acute COVID-19: Ongoing symptomatic phase vs. post-COVID-19 syndrome"

**EMBASE**

'coronavirus disease 2019'/exp

((long OR 'long term' OR 'long lasting' OR Prolonged) NEAR/3 ('2019 ncov' OR ncovor OR covid OR 'sars cov 2')):ti,ab

(('long* haul*' OR longhaul* OR 'long* tail*' OR longtail* OR longduration* OR 'long duration*' OR longlast* OR 'long last*') NEAR/3 ('2019 ncov' OR ncovor OR covid OR 'sars cov 2')):ti,ab

((chronic OR post-infect* OR 'post acute' OR persistent) NEAR/6 ('2019 ncov' OR ncovor OR covid OR 'sars cov 2')):ti,ab

('post covid' OR postcovid OR ‘Post acute covid*’ OR ‘postacute covid*’):ti,ab

#1 OR #2 OR #3 OR #4 OR #5

'hospital discharge'/exp

((long OR 'long term' OR 'long lasting' OR post-acute OR post-covid OR Postdischarge OR Post-discharge OR chronic OR persist* OR linger*) NEAR/3 (effect* OR symptom* OR recover* OR therapy OR sequelae OR complication* OR consequence* OR syndrome* OR outcome*)):ti,ab

((prolong* OR longer OR extended OR lengthy OR lengthen OR increase* OR drawn-out OR proctracted) NEAR/4 recover*):ti,ab

#7 OR #8 OR #9

'fatigue'/de OR 'exhaustion'/de OR 'fatigue assessment scale'/exp OR 'fatigue assessment instrument'/exp OR 'postviral fatigue syndrome'/exp OR 'Fatigue Severity Scale'/exp OR 'Fatigue Impact Scale'/exp

(fatigue* OR tired* OR exhaustion OR exhausted OR lethargy OR sluggish OR 'postviral syndrome' OR 'post viral syndrome' OR 'fatigue severity scale' OR ‘myalgic encephalomyelitis’):ti,ab

#11 OR #12

#6 AND #10 AND #13

---

**Medline**

COVID-19/

((long OR long term OR long lasting OR Prolonged) adj3 (2019 ncov OR ncovor OR covid OR sars cov 2)).ti,ab.

((long* haul* OR longhaul* OR long* tail* OR longtail* OR longduration* OR long duration* OR longlast* OR long last*) adj3 (2019 ncov OR ncovor OR covid OR sars cov 2)).ti,ab.

((chronic OR post infect* OR post acute OR persistent) adj6 (2019 ncov OR ncovor OR covid OR sars cov 2)).ti,ab.

(post covid OR postcovid OR Post acute covid* OR postacute covid*).ti,ab.

or/1-5

Patient Discharge/

((long OR long term OR long lasting OR post acute OR post covid OR Postdischarge OR Post discharge OR chronic OR persist* OR linger*) adj3 (effect* OR symptom* OR recover* OR therapy OR sequelae OR complication* OR consequence* OR syndrome* OR outcome*)).ti,ab.

((prolong* OR longer OR extended OR lengthy OR lengthen OR increase* OR drawn out OR proctracted) adj4 recover*).ti,ab.

or/7-9

Fatigue/ OR Fatigue Syndrome, Chronic/ OR Muscle Weakness/

(fatigue assessment scale OR fatigue assessment instrument OR Fatigue Severity Scale OR Fatigue Impact Scale).ti,ab.

(fatigue* OR tired* OR exhaustion OR exhausted OR lethargy OR sluggish OR postviral syndrome OR post viral syndrome OR fatigue severity scale OR myalgic encephalomyelitis).ti,ab.

or/11-13

6 AND 10 AND 14

---

**ProQuest Coronavirus Research Database**

TI ((long OR “long term” OR “long lasting” OR Prolonged) N2 (“2019 ncov” OR ncovor OR covid OR “sars cov 2”)) OR AB ((long OR “long term” OR “long lasting” OR Prolonged) N3 (“2019 ncov” OR ncovor OR covid OR “sars cov 2”))

TI ((“long* haul*” OR longhaul* OR “long* tail*” OR longtail* OR longduration* OR “long duration*” OR longlast* OR “long last*”) N2 (“2019 ncov” OR ncovor OR covid OR “sars cov 2”)) OR AB ((“long* haul*” OR longhaul* OR “long* tail*” OR longtail* OR longduration* OR “long duration*” OR longlast* OR “long last*”) N2 (“2019 ncov” OR ncovor OR covid OR “sars cov 2”))

TI ((chronic OR post-infect* OR “post acute” OR persistent) N5 (“2019 ncov” OR ncovor OR covid OR “sars cov 2”)) OR AB ((chronic OR post-infect* OR “post acute” OR persistent) N5 (“2019 ncov” OR ncovor OR covid OR “sars cov 2”))

TI (“post covid” OR postcovid OR “Post acute covid*” OR “postacute covid*”) OR AB (“post covid” OR postcovid OR “Post acute covid*” OR “postacute covid*”)

1 OR 2 OR 3 OR 4

TI (“fatigue assessment scale” OR “fatigue assessment instrument” OR “postviral fatigue syndrome” OR “Fatigue Severity Scale” OR “Fatigue Impact Scale”) OR AB (“fatigue assessment scale” OR “fatigue assessment instrument” OR “postviral fatigue syndrome” OR “Fatigue Severity Scale” OR “Fatigue Impact Scale”)

TI (fatigue* OR tired* OR exhaustion OR exhausted OR lethargy OR sluggish OR “postviral syndrome” OR “post viral syndrome” OR “fatigue severity scale” OR “myalgic encephalomyelitis”) OR AB (fatigue* OR tired* OR exhaustion OR exhausted OR lethargy OR sluggish OR “postviral syndrome” OR “post viral syndrome” OR “fatigue severity scale” OR “myalgic encephalomyelitis”)

6 OR 7

S5 AND S8

---

**Google Scholar**

2019nCoV|"2019 nCoV|CoV|coronavirus"|"2019 novel|new coronavirus|cov"|"wuhan coronavirus|cov|ncov|outbreak"|"wuhan*coronavirus|cov|ncov|outbreak"|"wuhan**coronavirus |cov|ncov|outbreak"|"coronavirus|cov|ncov*wuhan“ fatigue|exhaustion

---

**LitCOVID**

(“long term”[tiab] OR “long lasting”[tiab] OR Prolonged[tiab] OR chronic[tiab] OR post-infect*[tiab] OR “post acute”[tiab]) AND (fatigue*[tiab] OR tired*[tiab] OR exhaustion[tiab] OR exhausted[tiab] OR lethargy[tiab] OR “postviral syndrome”[tiab] OR “post viral syndrome”[tiab] OR “myalgic encephalomyelitis”[tiab])
