## Appendix B for "A systematic review of persistent symptoms and residual abnormal functioning following acute COVID-19: Ongoing symptomatic phase vs. post-COVID-19 syndrome"

| First Author | Date | Country of Authorship | Primary Topic | Specific Topic(s) | Study Design (re. long COVID) | N | Age (Years) | Gender (% Female) | Eligibility Criteria* | COVID Hospital Status | Time Post-COVID (from recovery/onset) | Weeks from COVID Onset | Fatigue Measurement Tool (FMT)* | Method of FMT Administration |
|---|---|---|---|---|---|---|---|---|---|---|---|---|---|---|
| **Arnold** | Apr-21 | United Kingdom | Symptomology | 1) Symptomatology; 2) Respiratory functioning<br>3) Quality of life; 4) Haemotology | Cross-Sectional | 110 | **M** = 60<br>**IQR** = 46-73 | 38% | - | Inpatient | **M** = 83 days<br>**IQR** = 74-88 | 16 weeks | SF-36 GSQ | Summed rating scale<br>in-person |
| **Bellan** | Jan-21 | Italy | Respiratory | 1) Respiratory functioning<br>2) Symptomatology | Cross-Sectional | 238 | **M** = 61<br>**IQR** = 50-71 | 40% | - | Inpatient (+ICU) | **All** ~ 4 months | 21 weeks | Novel GSQ | Binary scale (Y/N)<br>via phone |
| **Carfi** | Jul-20 | Italy | Symptomology | 1) Symptomatology | Short Study | 143 | $\bar{X}$ = 56.5<br>**SD** = 14.6 | 37% | - | Inpatient (+ICU) | $\bar{X}$ = 36.1 days<br>**SD** = 12.9 | 9 weeks | Novel GSQ | Binary scale (Y/N)<br>in-person |
| **Carvalho-Schneider** | Oct-20 | France | Symptomology | 1) Symptomatology | Longitudinal | 150 | $\bar{X}$ = 49.0<br>**SD** = 15.0 | 56% | **Excl**: ICU or residential care | Mixed (-ICU) | **TI**: $\bar{X}$ = 32.7 days, **SD** = 2.5*<br>**TII**: $\bar{X}$= 59.7 days, **SD** = 1.7* | 5 weeks \| 9 weeks | - | - |
| **Cheng** | Jan-21 | United Kingdom | Symptomology | 1) Symptomatology<br>2) Respiratory functioning | Cross-Sectional | 113 | **M** = 73<br>**IQR** = 57-84 | 44% | - | Inpatient (+ICU) | **R** = 6-12 weeks | 13 weeks | Novel GSQ | Binary scale (Y/N)<br>in-person |
| **Chopra** | Nov-20 | USA | Symptomology | 1) Symptomatology | Short Study | 488 | **M** = 62<br>**IQR** = 50-72 | 48% | - | Inpatient (+ICU) | **All** ~ 60 days | 13 weeks | - | - |
| **Cortés-Telles** | Jun-20 | Mexico | Symptomology | 1) Symptomatology<br>2) Respiratory functioning | Cross-Sectional | 186 | $\bar{X}$ = 47.0<br>**SD** = 13.0 | 39% | **Excl**: Critical cases | Mixed | $\bar{X}$ = 59.5 days*<br>**SD** = 13.5 | 9 weeks | Novel GSQ | Binary scale (Y/N)<br>in-person |
| **Daher** | Oct-20 | Germany | Respiratory | 1) Symptomatology; 2) Respiratory functioning<br>3) Cognitive/Psych func.; 4) Inflamm. response | Cross-Sectional | 33 | $\bar{X}$ = 64.0<br>**SD** = 3.0 | 33% | **Incl**: Severe COVID diagnosis<br>**Excl**: ARDS diagnosis & ICU | Inpatient | **M** = 56 days<br>**IQR** = 48-71 | 12 weeks | Novel GSQ | Binary scale (Y/N)<br>in-person |
| **D'Cruz** | Jan-21 | United Kingdom | Symptomology | 1) Symptomatology<br>2) Respiratory functioning | Cross-Sectional | 119 | $\bar{X}$ = 58.7<br>**SD** = 14.4 | 38% | - | Inpatient (+ICU) | **M** = 61 days<br>**IQR** = 51-67) | 13 weeks | Novel GSQ | Binary scale (Y/N)<br>in-person |
| **De Lorenzo** | Oct-20 | Italy | Symptomology | 1) Symptomatology | Cross-Sectional | 185 | **M** = 57<br>**IQR** = 48-67 | 34% | - | Mixed | **M** = 23 days<br>**IQR** = 20-29 | 7 weeks | - | - |
| **Froidure** | Apr-21 | Belgium | Respiratory | 1) Respiratory functioning<br>2) Symptomatology | Cross-Sectional | 134 | **M** = 60<br>**IQR** = 53-68 | 41% | **Excl**: Follow-ups 60<x<120 days | Inpatient (+ICU) | **M** = 95 days<br>**IQR** = 86-107 | 18 weeks | Novel GSQ | Binary scale (Y/N)<br>in-person |
| **Garrigues** | Aug-20 | France | Symptomology | 1) Symptomatology | Short Study | 120 | $\bar{X}$ = 63.2<br>**SD** = 15.7 | 37% | - | **Multiple Cohorts**:<br>Inpatient \| ICU | $\bar{X}$ = 110.9 days*<br>**SD** = 11.1 | 16 weeks | Novel GSQ | Binary scale (Y/N)<br>via phone |
| **Halpin** | Feb-21 | United Kingdom | Symptomology | 1) Symptomatology; 2) Rehabilitation needs<br>3) Psych. wellbeing; 4) Quality of life; 5) Other | Cross-Sectional | 100 | **M** = 70.5 / 58.5<br>**R** = 20-93 / 34-84 | 46% | **Incl**: Discharged<br>**Excl**: Comorbid neuro. condition | **Multiple Cohorts**:<br>Inpatient \| ICU | $\bar{X}$ = 48.0 days<br>R = 29-71 | 11 weeks | Novel GSQ | Likert scale<br>via phone |
| **Huang (I)** | Jan-21 | China | Symptomology | 1) Symptomatology; 2) Respiratory functioning<br>3) Quality of life; 4) Bloods | Cross-Sectional | 1,733 | **M** = 57<br>**IQR** = 47-65 | 48% | **Excl**: Comorbid psych. Condition; Re-admission; Immobility issues | Inpatient (+ICU) | **M** = 153 days<br>**IQR** = 146-160 | 26 weeks | Novel GSQ | Binary scale (Y/N)<br>in-person |
| **Huang (II)** | Jun-20 | China | Respiratory | 1) Respiratory functioning | Cross-Sectional | 57 | $\bar{X}$ = 46.7<br>**SD** = 13.8 | 54% | **Excl**: Previous pulmonary resection, neurological disease, or mental illness | Inpatient | **All** = 30 days | 8 weeks | - | - |
| **Iqbal** | Feb-21 | Pakistan | Symptomology | 1) Symptomatology | Cross-Sectional | 158 | $\bar{X}$ = 32.1<br>**SD** = 12.4 | 55% | **Excl**: Psychiatric illness | Mixed | $\bar{X}$ = 38.1 days<br>**SD** = 20.0 | 7 weeks | Novel GSQ | Binary scale (Y/N)<br>via phone |
| **Jacobs** | Dec-20 | USA | Symptomology | 1) Symptomatology<br>2) Quality of life | Longitudinal | 183 | **M** = 57<br>**IQR** = 48-68 | 38% | **Incl**: Hospital for 3+ days<br>**Excl**: Dementia/Delirium | Inpatient | **TI** = 0 days \| **T2** = 14 days \| **TIII** = 21 days \| **TIV** = 35 days (IQR=30-40) | 4 weeks \| 6 weeks \|<br>7 weeks \| 9 weeks | PROMIS GSQ | Likert scale<br>via phone/email |
| **Lerum** | Apr-21 | Norway | Respiratory | 1) Respiratory functioning | Cross-Sectional | 103 | **M** = 59<br>**IQR** = 49-72 | 48% | - | **Multiple Cohorts**:<br>Inpatient \| ICU | **M** = 83 days*<br>**IQR** = 73-90 | 12 weeks | - | - |
| **Liang** | Oct-20 | China | Symptomology | 1) Symptomatology<br>2) Respiratory functioning | Cross-Sectional | 76 | **M** = 41.3<br>**IQR** = 13.8 | 72% | **Excl.** Comorbid neuro. condition; Underwent pulmonary resection | Inpatient (+ICU) | **Ti** = 1 month, **TII** = 2 months, **TIII** = 3 months; **SD** = 1 | 5 weeks \| 13 weeks \| 17 weeks | Novel GSQ | Binary scale (Y/N)<br>in-person |
| **Loerinc** | Mar-21 | USA | Other | 1) Transition of COVID care at discharge<br>2) Symptomatology | Cross-Sectional | 310 | **M** = 58<br>**Range** = 23-99 | 51% | - | Inpatient (+ICU) | **All** = Discharge | 4 weeks | Novel GSQ | Obtained from physician discharge notes |
| **Mandal** | Sep-20 | United Kingdom | Symptomology | 1) Long COVID<br>2) Respiratory functioning | Cross-Sectional | 384 | $\bar{X}$ = 59.9<br>**SD** = 16.1 | 38% | - | Inpatient (+ICU) | **M** = 54 days<br>**IQR** = 47=59 | 12 weeks | Novel GSQ | Likert scale<br>via phone / in-person |
| **Miyazato** | Oct-20 | Japan | Symptomology | 1) Symptomatology | Longitudinal | 63 | $\bar{X}$ = 48.1<br>**SD** = 18.5 | 33% | - | Inpatient | $\bar{X}$ = 108.0 days, **SD** = 23.0<br>(**TI** ~ 60 days, **TII** ~ 120 days)* | 9 weeks \| 17 weeks | Novel GSQ | Binary scale (Y/N)<br>via phone |
| **Mo** | Jun-20 | China | Respiratory | 1) Respiratory functioning | Short Study | 110 | $\bar{X}$ = 49.1<br>**SD** = 4.0 | 50% | **Excl**: Critical cases | Inpatient | **All** = Discharge | 4 weeks | - | - |
| **Moreno-Pérez** | Mar-21 | Spain | Symptomology | 1) Symptomatology<br>2) Respiratory functioning | Cross-Sectional | 277 | **M** = 62<br>**IQR** = 53-72 | 47% | - | Mixed | **M** = 77 days*<br>**IQR** = 71-85 | 11 weeks | Novel GSQ | Binary scale (Y/N)<br>in-person |
| **Osikomaiya** | Mar-21 | Nigeria | Symptomology | 1) Symptomatology | Cross-Sectional | 274 | $\bar{X}$ = 41.8<br>**SD** = 11.8 | 34% | - | Outpatient | **M** = 15 days<br>**IQR** = 14-17 | 6 weeks | Novel GSQ | Binary scale (Y/N)<br>in-person |
| **Prieto** | Mar-21 | Argentina | Symptomology | 1) Symptomatology | Cross-Sectional | 85 | $\bar{X}$ = 43.0<br>**SD** = 13.0 | 45% | - | Mixed | **M** = 53 days*<br>**IQR** = 31-65 | 8 weeks | Novel GSQ | Binary scale (Y/N)<br>in-person |
| **Raman** | Nov-20 | United Kingdom | Symptomology | 1) Symptomatology; 2) Neurological functioning<br>3) Respiratory func.; 4) Psych. Wellbeing; Other | Cross-Sectional | 58 | $\bar{X}$ = 55.4<br>**SD** = 13.2 | 41% | **Incl**: Moderate-to-severe COVID | Inpatient (+ICU) | **M** = 2.3 months*<br>**IQR** = 2.06-2.53 | 10 weeks | Fatigue Severity Scale | Likert scale<br>in-person |
| **Rosales-Castillo** | Jan-21 | Spain | Symptomology | 1) Symptomatology<br>2) Psychological wellbeing | Cross-Sectional | 118 | $\bar{X}$ = 60.2<br>**SD** = 15.1 | 44% | - | Inpatient | $\bar{X}$ = 50.8 days<br>**SD** = 6.0 | 11 weeks | Novel GSQ | Binary scale (Y/N)<br>in-person |
| **Shah** | Mar-21 | Canada | Respiratory | 1) Respiratory functioning<br>2) Symptomatology | Cross-Sectional | 60 | **M** = 67<br>**IQR** = 54-74 | 32% | - | Inpatient | $\bar{X}$ = 11.7 weeks*<br>**SD** = n/a | 12 weeks | - | - |
| **Simani** | Feb-21 | Iran | Symptomology | 1) Symptomatology<br>2) Psychological wellbeing | Cross-Sectional | 120 | $\bar{X}$ = 54.6<br>**SD** = 16.9 | 33% | **Excl**: Previous life-threatening infection | Inpatient (+ICU) | **All** ~ 6 months | 30 weeks | Fukuda questionnaire | Likert scale<br>via phone |
| **Sykes** | Apr-21 | United Kingdom | Symptomology | 1) Symptomatology; 2) Haemotology<br>3) Respiratory functioning | Cross-Sectional | 134 | **M** = 58<br>**R** = 25-89 | 34% | **Excl**: Mild symptoms or frail | **Multiple Cohorts**:<br>Inpatient \| ICU | M = 113 days, R = 46-167<br>(TI~61, TII~88, TIII~113, TIV~147) | 13 weeks \| 17 weeks\|<br>20 weeks \| 25 weeks | Novel GSQ | Binary scale (Y/N)<br>in-person |
| **Taboada** | Dec-20 | Spain | Quality of Life | 1) Quality of life<br>2) Symptomatology | Cross-Sectional | 91 | $\bar{X}$ = 65.5<br>**SD** = 10.4 | 35% | - | ICU | **All** ~ 6 months | 30 weeks | - | - |
| **Townsend (I)** | Nov-20 | Ireland | Symptomology | 1) Symptomatology<br>2) Haemotology | Cross-Sectional | 128 | $\bar{X}$ = 49.5<br>**SD** = 15.0 | 54% | **Incl**: 6+ weeks post-COVID | Mixed | **M** = 72 days<br>**IQR** = 62-87 | 14 weeks | Chalder Fatigue Scale<br>(incl. Psych/Phys sub-scales) | Likert scale<br>in-person |
| **Venturelli** | Jan-21 | Italy | Respiratory | 1) Symptomatology | Cross-Sectional | 767 | $\bar{X}$ = 63.0<br>**SD** = 13.6 | 33% | - | Inpatient (+ICU) | **M** = 105 days*<br>**IQR** = 84-127 | 15 weeks | Novel GSQ | Binary scale (Y/N)<br>in-person |
| **Walle-Hansen** | Mar-21 | Norway | Quality of Life | 1) Quality of life | Cross-Sectional | 106 | $\bar{X}$ = 74.3<br>**R** = 60-96 | 43% | - | Inpatient (+ICU) | **M** = 186<br>**IQR** = n/a | 31 weeks | - | - |
| **Wang** | May-20 | China | Symptomology | 1) Symptomatology; 2) Respiratory functioning<br>3) Haemotology | Longitudinal | 131 | **M** = 49<br>**IQR** = 36-62 | 55% | **Incl**: Objective improvements (incl. fever, chest CT, respiratory symptoms) | Inpatient | **TI** = 0 days, **TII** = 7-14 days<br>**TIII** = 21-28 days | 4 weeks \| 6 weeks \|<br>8 weeks | Novel GSQ | Binary scale (Y/N)<br>in-person |
| **Wong** | Nov-20 | Canada | Symptomology | 1) Symptomatology | Short Study | 78 | $\bar{X}$ = 62.0<br>**SD** = 16.0 | 36% | - | Inpatient | **M** = 13 weeks*<br>**IQR** = 11-14 | 13 weeks | - | - |
| **Xiong** | Sep-20 | China | Symptomology | 1) Symptomatology<br>2) Psychological wellbeing | Cross-Sectional | 538 | **M** = 52<br>**IQR** = 41-62 | 55% | **Incl**: 3+ months post-COVID | Inpatient | **M** = 97 days<br>**IQR** = 95-102 | 18 weeks | Novel GSQ | Binary scale (Y/N)<br>via phone |
| **Yu** | Mar-20 | China | Respiratory | 1) Respiratory functioning | Cross-Sectional | 32 | **M** = 44.4<br>**IQR** = N/A | 31% | **Incl**: 3 chest CT scans (2-hospital; 1-post) | Inpatient (+ICU) | **M** = 9 days<br>**IQR** = N/A | 5 weeks | Novel GSQ | Binary scale (Y/N)<br>in-person |

**excl. COVID+ test result, age & hospital status*

** = from symptom onset or hospitalisation*

*[blank] = from hospital discharge or COVID recovery*

**"From COVID onset" = +28 days until "recovery"*

*Novel GSQ = Novel general symptom questionnaire*
